# Changes in Female Cancer Diagnostic Billing Rates over the COVID-19 Period in the Ontario Health Insurance Plan

**DOI:** 10.1101/2024.12.17.24319153

**Authors:** Deanna McLeod, Ilidio Martins, Anna V. Tinker, Amanda Selk, Christine Brezden-Masley, Nathalie LeVasseur, Alon D. Altman

## Abstract

**Background and Objectives:** The initial response to COVID-19 in Ontario included suspension of cancer screening programs and deferral of diagnostic procedures and many treatments. Although the short-term impact of these measures on female cancers is well documented, few studies have assessed the mid- to long-term impacts. We conducted an analysis of Ontario Health Insurance Program (OHIP) claims for female cancer diagnostic codes by comparing annual billing prevalence and incidence rates during the COVID-19 period (2020-2022) to pre-COVID-19 levels (2015-2019).

**Methods:** Linear regression analysis was used to fit pre-COVID-19 (2015-2019) data for each OHIP billing code and extrapolate counterfactual values for the years of 2020-2022. Excess billing rates were calculated as the difference between projected and actual rates for each year.

**Results:** In 2020, OHIP billing prevalence rates for cervical, breast, uterine and ovarian cancer decreased relative to projected values for that year by -50.7/100k, -13.9/100k, -3.5/100k and -3.8/100k, respectively. The reverse was observed in 2021 with rate increases of 47.8/100k, 59.1/100k, 2.5/100k and 3.7/100k, respectively. In 2022, the excesses were further amplified, especially for cervical and breast cancers (111.2/100k and 78.67/100k, respectively). The net excess patient billing rate for 2020-2022 was largely positive for all female cancers (108.3/100k, 123.7/100k, 5.2/100k, 1.8/100k, respectively). Analysis of billing incidence rates showed similar trends.

**Conclusion:** The expected female cancer billing rate decreases in 2020 were followed by large increases in 2021 and 2022, resulting in a cumulative excess during the COVID-19 period. Further research is required to assess the nature of these changes.

## Introduction

The World Health Organization declared the coronavirus disease 2019 (COVID-19) a Public Health Emergency of International Concern on January 30, 2020 and recommendations were issued to prevent disease transmission.^1^ In the Canadian province of Ontario, a State of Emergency (SOE) was declared on March 17, 2020 leading to the implementation of public health measures such as social distancing, quarantine and isolation, restrictions and changes to in-person public health services and workforce as well as widespread closures of businesses and schools.^2^ As the first provincial SOE period expired on July 24, 2020, many public health measures were canceled or eased with others remaining in effect outside of the formal SOE period. Subsequent provincial SOEs were in place from January 14, 2021 to February 19, 2021 and April 8, 2021 to June 9, 2021. As in other jurisdictions, the longest SOE period(s) and the strictest restrictions occurred in 2020.^2,3^ Public health measures began to consistently ease in 2021 as levels of two-dose messenger ribonucleic acid (mRNA) vaccine coverage increased to close to 80% of eligible Ontarians by the end of 2021.^4^

Early public health measures had a considerable impact on cancer care delivery in Ontario including suspension of cancer screening programs and deferral of selected diagnostic and treatment procedures.^2,5^ Deferred services were permitted to resume gradually in Ontario beginning on May 26, 2020.^6^ However, many factors known to negatively impact access to cancer care continued, including general restrictions on mobility, patient volume limitations, reductions in the number of hospital beds for non-emergent conditions, and fear of COVID-19.^2,7^ This resulted in reductions in cancer screening and diagnoses^5,8^ in 2020 relative to pre-COVID-19 levels especially within but also outside SOE periods. Access to key services returned toward historical levels by the end of 2020 and early in 2021. Similar impacts were observed throughout Canada and other countries.^9–28^

Modeling studies have estimated the impact of delayed diagnosis and treatment on cancer outcomes,^9,12,15,22,29,30^ and the short-term consequences have been evaluated in both prospective and retrospective cohort studies.^14,18,20–22,31^ In particular, multiple studies have shown changes impacting female cancers, both gynecological and breast cancers, including: i) decreases in screening volumes and diagnoses, especially in screening-dependent cancers (breast and cervical) during 2020^14,23,25,32–47^ followed by increased diagnosis in 2021;^45,48^ ii) higher proportion of more advanced disease and stage migration;^33,34,37,44,47^ iii) curative-intent treatment delays;^36,38,42,49–53^ iv) changes to treatment protocols with a priority towards conservative treatment and neoadjuvant systemic therapies;^27,36,38,42,52,54–58^ and v) worse female cancer outcomes.^21,34,51^ However, the mid- to long-term impact of public health measures have yet to be evaluated for female cancers.

Cancer registries are a reliable and rich source of epidemiological data; however, they are associated with reporting delays which can extend 3 to 4 years or longer due to registration, processing and publication delays.^59–61^ Conversely, claims data are more timely and provide a source of real-world epidemiological data that has been extensively used for healthcare and cancer research.^62–65^ The Ontario Health Insurance Plan (OHIP) database collects insurance claims submitted to Ontario’s medical claims electronic transfer system and captures Health Care Payments rendered by physicians and private medical labs in Ontario’s public healthcare system, which services its 15 million residents rendering it a rich repository of epidemiological data. Our analysis seeks to assess the relative magnitude of changes in OHIP female cancer billings during the COVID-19 period (2020-2022) by comparison to a historical pre-COVID-19 reference period (2015-2019).

## Methods

The OHIP data used in our analysis was obtained from two Freedom of Information (FOI) requests submitted to the Ministry of Health Ontario. The first request included data from 2015 to the first quarter of 2022 and the second for the full year of 2022. Yearly unique patient counts (hereafter referred to as billing prevalence) and new patient counts (hereafter referred to as billing incidence) were provided for requested diagnostic codes. Prevalence data was collected by counting the first time a diagnostic code was billed for a patient within a given year while incidence data was collected by counting the first time a diagnostic code was billed for a patient within the total study period. Data provided were of unique patients as each patient was counted once per category per calendar year, leading to the exclusion of counts for additional visits of the same patient under the same diagnostic code. In order to account for changes in population over time, yearly OHIP billing rates were calculated by dividing patient counts by the total population of Ontario for that year as determined by Statistics Canada.^66^ Given challenges in interpreting incidence rates due to the varying lengths of reference or “look-back” periods,^67,68^ our analysis primarily focused on prevalence rates.

We compared yearly OHIP billing patterns from the COVID-19 period (2020-2022) to a historical pre-COVID-19 reference period (2015-2019). We used a linear regression analysis to fit pre-COVID-19 (2015-2019) data for each billing code and used the resulting linear relationship to extrapolate hypothetical values for a scenario where the COVID-19 crisis did not occur from 2020 to 2022 (hereafter referred to as predicted or projected). An F-test was used to assess whether a regression coefficient was required to fit the data, i.e. whether a linear model in which rates increased or decreased over time was significantly better in fitting a model in which they were constant. If the F-test failed, projected values for 2020-2022 were calculated as the average of pre-COVID-19 rates. Models of increased complexity (non-linear) were not used to fit the data. The 95% confidence intervals of the projected mean for COVID-19 years (2020-2022) are shown in billing rate plots (Figure 1). We calculated excess billing rates as the difference between expected and actual rates for each year.^69,70^ In order to provide a measure of the degree of departure from predictions, deviations from predicted values during the COVID-19 period were normalized using the standard deviation of the excess rates during the pre-COVID-19 reference period (standard score).^69^ For reference, an absolute standard score equal to or above the critical value of 1.96 falls in the rejection region of the hypothesis that the observable differences belong to the reference population at a confidence level of 95% under the assumption of a normal distribution.

**Figure 1.**
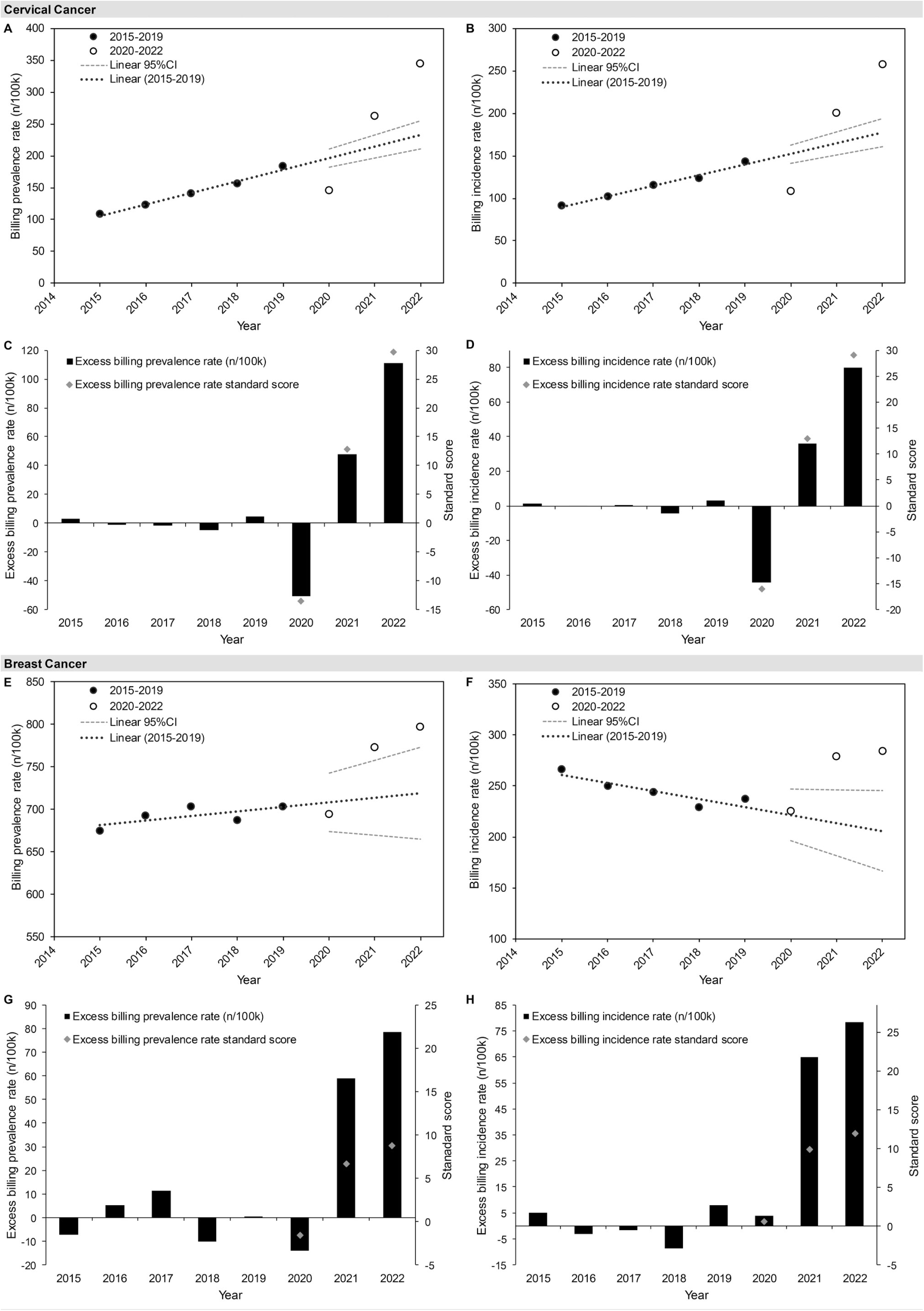
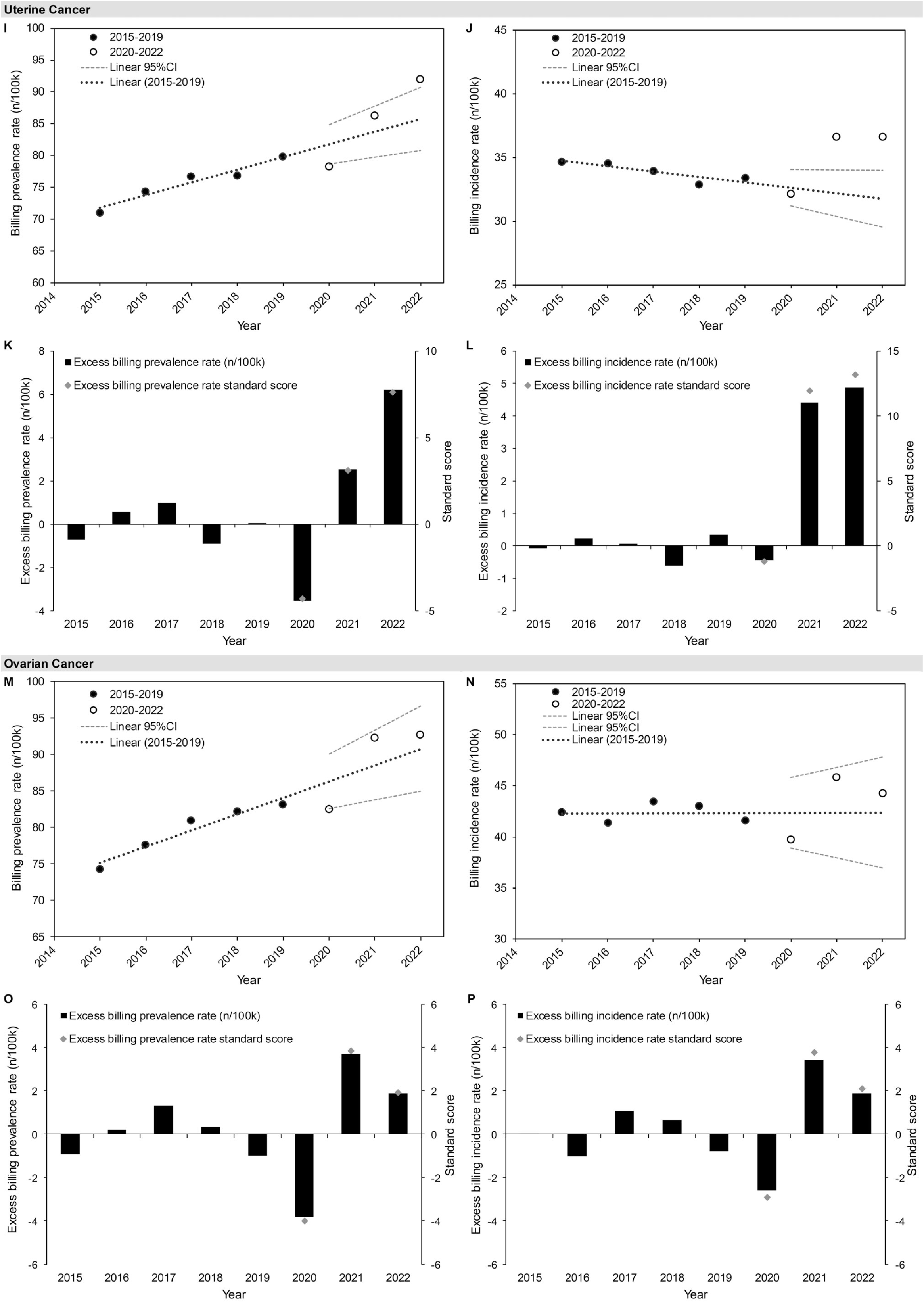
Yearly OHIP 2015-2022 billing prevalence and incidence rates and the respective excess rates and standard scoresa for cervical (a-d), breast (e-h), uterine (i-l) and ovarian (m-p) cancer diagnostic codes. Linear regression fits to billing rates from pre-COVID-19 years (2015-2019) and the 95% CIs of the projected mean for COVID-19 years (2020-2022) are shown in billing rate plots.

## Results

The fits of pre-COVID-19 data to linear regression models were significantly improved (F-test p-value <0.05) with the addition of regression coefficients in all datasets except for ovarian cancer incidence rates (p=0.053) which was therefore fit to a model where billing rates were constant over time. The differences between projected and observed billing rates (excess billing rates) for each COVID-19 year (2020-2022) rather than the respective billing rate values are described below.

### Cervical Cancer

Over the pre-COVID-19 reference period, OHIP billing prevalence rates for cervical cancer diagnostic code (180) increased steadily at a rate of 18.2/100k per year (Figure 1a). In 2020, billing prevalence rates dropped sharply with decreases of -50.7/100k relative to projected values, a change that was associated with a very large standard score (−13.6; Figure 1c). In 2021, rates reversed, rising by 47.8/100k relative to projected values (standard score 12.8), a change closely mirroring the previous year’s drop. In 2022, rates increased relative to expected values, nearly doubling compared to 2021 (111.2/100k; standard score 29.7). This resulted in a large positive net excess billing prevalence rate of 108.3/100k for the COVID-19 period. A similar trend that was also observed for OHIP billing incidence rates (Figure 1b/d).

### Breast Cancer

Prior to COVID-19, billing prevalence rates for breast cancer diagnostic code (174) oscillated between 674/100k and 703/100k with an overall increase of 7.8/100k per year (Figure 1e). In 2020, rates dropped by -13.9/100k relative to projected values, a change in line with the variability over reference period (standard score -1.6; Figure 1e/g). In 2021 and 2022, rates increased dramatically relative to projected values (59.1/100k and 78.6/100k, respectively), a change that was associated with large standard scores (6.6 and 8.8, respectively). The net excess billing prevalence rate for the COVID-19 period was largely positive (123.7/100k) pointing to a larger than expected increase in the prevalence of breast cancer billings. A similar pattern was seen for billing incidence rates except in the backdrop of downward trend over the reference period (Figure 1f/h).

### Uterine Cancer

Over the reference period, billing prevalence rates for uterine cancer diagnostic code (182) increased steadily by an average of 1.7/100k per year (Figure 1i). In 2020, rates decreased relative to projected values by -3.5/100k and were associated with large standard scores (−4.3; Figure 1k). A comparable and opposite change occurred in 2021 (2.5/100k, standard score 3.1) followed by a second large increase in 2022 (6.2/100k, standard score 7.6). The net excess billing prevalence rate for the COVID-19 period was positive (5.2/100k) indicating a higher-than-expected prevalence of uterine cancer billings. Billing incidence rates showed similar results albeit relative to a downward pre-pandemic trend, with smaller rate decreases in 2020 (−0.45, standard score -1.2) and larger increases in 2021 and 2022 that were associated with very large standard scores (11.9 and 13.1, respectively; Figure 1j/l).

### Ovarian Cancer

In the pre-COVID-19 reference period, billing prevalence rates for ovarian cancer diagnostic code (183) increased by an average of 1.8/100k per year (Figure 1m). In 2020, rates dropped relative to projected values (−3.8/100k) and were associated with large standard scores (−4.0; Figure 1o). Rates shifted in the opposite direction in 2021 (3.7/100k, standard score 3.9) and increased again in 2022 (1.9/100k, standard score 1.9). The net excess prevalence rate for the COVID-19 period was positive (1.8/100k) reflecting a slightly higher than expected rate of ovarian cancer billings. OHIP incidence rates through the COVID-19 period were similar to those of the historical reference period which remained constant over time (Figure 1n/p).

## Discussion

### What were our main findings?

Our analysis of OHIP claims data identified substantial changes in billing rates for female cancers over the COVID-19 period (2020-2022) compared to the 5-year pre-COVID-19 reference period. Most of these changes were large relative to projected values. This was apparent both from visual inspection of the plots and from the relative magnitude of their excess rate values which were often many times the standard deviation of the historical sample (Figure 1 and Table 1), suggesting that these changes are unlikely due to chance. In 2020, all female cancers with the exception of breast cancer showed large decreases in billing rates, consistent with previously described disruptions in screening programs and overall reductions in access to care due to public health measures that year.^8–22,71^ All female cancers showed an increase or “rebound” billing in 2021 consistent with women re-accessing care. The rebound was proportional to the decrease in 2020 for all female cancer except for breast cancer which was 4-fold higher. Billing rates continued to rise in 2022 resulting in net excess rates over the COVID-19 for all female cancers. These net excess rates were 2-fold and 9-fold higher than decreases seen in 2020 for cervical and breast cancer, respectively.

**Table 1.** OHIP excess billing prevalence rates and respective standard scores by year and net excess patient billing rates over the COVID-19 period

| OHIP Female Cancer Billing Code | 2020 |  | 2021 |  | 2022 |  | Net Excess (n/100k) |
| --- | --- | --- | --- | --- | --- | --- | --- |
|  | Excess (n/100k) | SS | Excess (n/100k) | SS | Excess (n/100k) | SS |  |
| **180 - Cervical** | -50.7 | -13.6 | 47.8 | 12.8 | 111.2 | 29.7 | 108.3 |
| **174 - Breast** | -13.9 | -1.6 | 59.1 | 6.6 | 78.6 | 8.8 | 123.7 |
| **182 - Uterine** | -3.5 | -4.3 | 2.5 | 3.1 | 6.2 | 7.6 | 5.2 |
| **183 - Ovarian** | -3.8 | -4.0 | 3.7 | 3.9 | 1.9 | 1.9 | 1.8 |

**Abbreviations:** n, number; OHIP, Ontario Health Insurance Plan; SS, standard score

### Could changes in billing patterns explain the changes in billing rates for female cancers?

OHIP captures the majority of medical encounters in Ontario’s public health system and only misses a fraction of acute care and emergency room visits.^72^ As approximately 25% of cancer cases present in acute settings^73–77^ and these encounters require confirmation of diagnosis or follow-up assessment with specialists in the community, our analysis likely represents a comprehensive and representative assessment of claims data for female cancers.

It is well recognized that claims data are susceptible to changes in billing practices in the form of billing errors, contract changes, implementation of new billing systems, and/or shifts in the nature of care delivered.^78–81^ Medical billing data is subject to biases and inaccuracies,^82–85^ especially when conducted by support staff such as students and nurses, and such billing errors may be more frequent in systems that are rarely audited such as OHIP.^86^ Assuming that billing inaccuracies related to physician coding remained constant over time, our use of a historical reference period should serve to control for the impact of these errors. Therefore, is unlikely that billing errors alone could explain the striking changes in billing rates for female cancers over the COVID-19 period.

As the health system in Ontario is publicly funded, the province is the principal employer and therefore contract changes could potentially impact billing patterns, especially if these changes occurred during the COVID crisis. On April 22, 2020 the Ontario government initiated a series of economic initiatives starting with the COVID–19 Advance Payment Program to help accommodate increased demand for care in light of pre-existing shortages in family physicians.^87^ On April 1, 2021 and April 1, 2022 Ontario also introduced pay-for-performance incentives offering a 1% increase for all physician services and a 2% increase for primary health care services, respectively.^88,89^ According to the National Physician Database, provincial efforts were met with some success. The 2.5% drop in total clinical payments to physicians from the 2020-2021 fiscal year was followed by increases of 9.2% and 6.4% in 2021-2022 and 2022-2023, respectively^90^ and physician coverage rates had recovered to pre-COVID-19 levels in 2021 (235/100k versus [vs.] 229/100k in 2020 vs. 234/100k in 2019).^91^ However, the coverage of overall physician services remained lower in 2020-2021 and 2021-2022 (714,806/100k and 722,911/100k, respectively) relative to both 2019-2020 and 2018-2019 (739,456/100k and 742,284/100k) fiscal years.^92^ Given the timing of the Ontario contract changes, it is possible that they affected billing rates for female cancer over the COVID-19 period, although their impact is likely limited since physician service coverage rates did not exceed pre-COVID-19 levels.

The implementation of new billing systems or shifts in referral patterns from hospitals to community settings could also impact billing patterns. Electronic health record (EHR) systems such as Epic, MEDITECH and Cerner, which seek to standardize billing could have a major impact on billing patterns. Ontario has been updating EHR hospital systems and networks in a stepwise fashion over the study period (2015-2022),^93^ however it is unclear to which degree changes in hospital billing would have on OHIP billing which predominately reflects community practice. To assess whether these or other system-wide changes in billing practices affected billing patterns, we examined the total number of unique patients (approximately 6 million) in the larger FOI-requested dataset representing both cancer and non-cancer billing codes to determine whether similar changes in billing rates were observed. We observed a considerable decrease in billing prevalence rates in 2020 (−11.3% relative to the 2015-2019 average; Figure S1) followed by a partial recovery in 2021 (6.5% relative to 2020), which remained lower than the reference period average (by 5.5%). It is therefore unlikely that system-wide changes in billing systems explain the dramatic changes in billing rates for female cancers over the COVID-19 period.

### Could shifts in the pattern of overall care explain the changes in female cancer billing rates?

Shifts in patterns of care which includes shifts to community care and virtual care can impact billing patterns. Early in the pandemic (March to July 2020), implementation of COVID-19 response measures resulted in a reduction in both the volume of patient care provided as well as surgical capacity.^94–96^ Surgical volumes returned to pre-pandemic levels by August 2020 and only temporary drops were observed at the peak of subsequent waves. A review of inpatient hospital occupancy through 2021 showed a recovery in occupancy to pre-COVID-19 levels by June 2020 followed by a further drop in occupancy beginning in March 2021 and extending to June 2021. It is therefore possible that reductions in hospital capacity could have prompted less intensive (non-surgical) patient care services to be transferred to the community contributing to the observed 2021 increase in OHIP physician billing rates.

A major component of the COVID-19 response in Ontario involved the expansion of telemedicine through the introduction of virtual visit K-codes.^97,98^ This facilitated a major transition from in-person to virtual care in 2020 resulting in a more than doubling of the number of physicians providing virtual care.^99^ Despite this, an overall decrease in the total number of visits and services occurred in both ambulatory and primary care settings during the first months of the COVID-19 period.^97,100–102^ As the healthcare system recovered in late in 2020, there was an accompanying return to in-person care by January 2021 with the total number of visits returning to pre-COVID-19 levels despite a greater proportion of virtual to in-person visits.^97,103,104^ Although a shift to virtual care occurred in Ontario over the COVID-19 period, it is unlikely to explain the increased billing rates for female cancers observed in 2021-2022 as the total number of visits returned to pre-COVID-19 levels in 2021. In addition, the provided OHIP dataset only records the first visit per person per condition within a given year, and the proportion of virtual to in-person visits during the COVID-19 period was highest in 2020 when billing prevalence rates were lowest, and lowest in 2022 when rates were highest.

### Can changes to Ontario screening volumes explain changes in billing patterns for female cancers?

The magnitude of net positive excess rates during the COVID-19 period was highest for screening-dependent cancers (cervical and breast), reaching values that were about one order of magnitude higher than those of other female cancers (uterine and ovarian). Ontario has well-established screening programs for breast (Ontario Breast Screening Program [OBSP] for 50-74 years-old and High-Risk OBSP for high-risk 30-69 years-old) and cervical (Ontario Cervical Screening Program for 21-69 years-old) cancers.^105^ To assess whether changes in screening volumes contributed to the changes in billing rates of screening-dependent female cancers, we examined changes in cervical and breast screening rates relative to OHIP findings. We found that screening volumes and OHIP billing rates followed a similar pattern of change through 2020 and 2021. Both cervical and breast monthly screening volumes were drastically reduced from March to July 2020 resulting in an overall reduction that year.^106,107^ In 2021-2022, cervical cytology screening volumes returned to or exceeded pre-pandemic levels with colposcopy volumes returning close to, yet consistently below, 2019 levels. Breast cancer screening volumes returned to or exceeded pre-pandemic levels through 2021 and 2022 with high-risk breast magnetic resonance imaging volumes recovering as early as August 2020 and staying above pre-pandemic levels for most of 2021-2022. Changes in screening volumes provide a compelling explanation for the observed fluctuations in OHIP billing rates for both cervical and breast cancer through 2020 and 2021.

Reductions in screening volumes in 2020 likely resulted in screening backlogs even after screening volumes returned to normal.^108,109^ In order to address these backlogs, Ontario implemented various “catch-up” strategies through 2020 and 2021 which prioritized screening based on level of cancer risk.^106,110^ A descriptive study of Ontario breast cancer screening found that this approach effectively cleared backlogs in the high-risk group (96.5%) by March 2021, a move that was associated with an 11% increase in the volume of abnormal results (relative to March 2019) despite a similar number of total screens.^110^ This strategy, however, was less effective at reducing backlogs in other risk groups. Backlogs in initial and follow-up screens for the average-risk group decreased by a mere (1% and 13.5% reductions, respectively) and backlogs in the low-risk group (biennial rescreens) increased by 7.6%.^110^ Given these findings, it is likely that prioritization strategies contributed to the rise in 2021 billing rates which could have extended into 2022. Prioritization strategies following deferred screening may also help explain the higher proportion of more advanced disease and stage migration,^33,34,37,44,47^ and worse female cancer outcomes observed in previous studies.^21,34,51^ It, however, is less likely to explain the large excess rates over the COVID-19 period that were 2-fold and 9-fold higher for cervical and breast cancer, respectively.

The changes in billing rates in 2020 and 2021 are generally consistent with changes in breast and cervical cancer screening volumes or diagnoses reported in the published literature. A systematic review of studies assessing the impact of public health measures on breast diagnoses found that over half the studies reported ≥49% reductions in screening volumes in 2020.^34^ An analysis of the United Kingdom primary care Clinical Practice Research Datalink GOLD database by Barclay et al. reported a decline in breast cancer diagnoses in 2020 followed by an increase in 2021.^45^ A study by Eijkelboom et al. reported a decrease in the monthly incidence of breast cancer in Norway and the Netherlands during the first wave (March-September 2020) followed by an increase in the incidence of stage IV breast cancer from August to December 2021.^33^ Finally, an analysis Knoll et al. reported a 52% decrease in newly diagnosed breast cancers during the 2020 lockdown periods followed by an increase in the proportion of symptomatic patients presenting upon re-opening compared to pre-COVID 2019 levels.^111^ Similar links between screening volumes and increased cancer incidence and severity were apparent for cervical cancer. Oymans et al. reported a decrease in incidence of cervical cancer in the Netherlands during the first COVID-19 wave (March-June 2020) followed by increases in incidence later in 2020 and in 2021.^32^ Analysis of the Japanese Society of Obstetricians and Gynecologic Oncology registry database found that incidence of cervical intraepithelial neoplasia 3 decreased in 2020 followed by an increase of similar magnitude in 2021; a significant shift towards higher tumor-node-metastasis stages of cervical cancer was observed in 2021.^48^ A single-center, retrospective study looking at cervical cancer screening volume and incidence over the pandemic period reported a higher percentage of patients presenting with cervical bleeding and a significant increase in more severe lesions despite lower colposcopy rates relative to the pre-pandemic period.^37^ A retrospective, single-center study of participants in a cervical cancer screening program found an absolute increase in cervical cancer incidence and a significant increase in the incidence of cervical intraepithelial neoplasia 2/3 lesions in the COVID-19 period relative to 2017-2019.^112^ Finally, a study of the Information System for Primary Care Services from the Public Health System of Catalonia, Spain found an increased prevalence of abnormal cervical cytology in 2020 and 2021 (ratio range=1.1–1.5) despite lower cervical cancer screening participation in those years (38.8% and 2.2%, respectively).^113^ Multiple studies show a relationship between screening volume changes and cancer incidence in 2020 and 2021.

### Could excess billing rates over the COVID-19 period reflect a rise in cancer rates?

Claims data has been used for epidemiological purposes. It differs, however, from cancer registry data in that the diagnostic code assigned to the billing occurs at the first medical encounter for a given cancer while registry data reflects a confirmed diagnosis established often after a series of tests and consultations. It is therefore possible that the OHIP billing prevalence numbers include encounters for suspected cancers or other events that may not reflect confirmed cancers. To assess this, we compared OHIP billing prevalence for female cancers to confirmed cancer prevalence figures reported by Cancer Care Ontario for the years of 2016 and 2018.^114,115^ We found that annual OHIP billing prevalence were around or below the upper limit of the thirty-year prevalence for all female cancers except for cervical cancer which was more than double (Table 2). This suggests that OHIP billing counts likely include both confirmed and suspected cancers and likely represent an overestimation of cancer rates.

**Table 2.** Comparison between 30-year prevalence counts reported by OH/CCO^114,115^ and yearly OHIP unique patient counts for cervical, breast, ovarian and uterine cancers for the years of 2018 and 2016

|  | Cervical Cancer | Breast Cancer | Ovarian Cancer | Uterine Cancer |
| --- | --- | --- | --- | --- |
| **2018** (Source: CCO – Ontario Cancer Statistics 2022) |  |  |  |  |
| Thirty-year prevalence count (n) | 10,306 | 141,829 | 10,277 | 32,438 |
| OHIP billing prevalence count (n) | 22,297 | 98,501 | 11,776 | 11,018 |
| **2016** (Source: CCO – Ontario Cancer Statistics 2020) |  |  |  |  |
| Thirty-year prevalence count (n) | 10,103 | 133,813 | 9,760 | 29,815 |
| OHIP billing prevalence count (n) | 17,022 | 96,087 | 10,763 | 10,322 |

**abbreviations:** CCO, Cancer Care Ontario; OH, Ontario Health; n, number; OHIP, Ontario Health Insurance Plan

### Could changes in cancer care contribute to changes in billing patterns?

Changes in patients presenting with either new, recurring, or worsening cancer symptoms or in the patterns of cancer care could contribute to the changes in billing rates for female cancers. It is well documented that the overall reduction in access to care and changes in oncological care to minimize exposure to COVID-19 during the first year of the pandemic resulted in decreases in surgical cancer treatment, biopsies, and new consultations and therapy suite visits for systemic and radiation treatments in Ontario.^5^ In addition, treatment protocols were adjusted to reduce hospital visits and time in cancer units, for instance incorporating neoadjuvant systemic therapy to minimize risk of surgical delays, switching to oral therapy, and use of monthly instead of weekly regimens of intravenous therapy.^5^ Treatment delays, reduced compliance with oral therapy and lack of adherence to management guidelines have been shown to negatively impact cancer outcomes such as quality of life, event-free survival and overall survival, particularly in female cancers.^29,116–127^ Disparities in time to cancer treatment have been previously identified showing longer wait times for women.^128–130^ It is therefore possible that changes in care, which occurred primarily in 2020 and 2021 may have contributed to female cancer billing rate changes over the COVID-19 period.

Multiple studies have reported shifts in cancer trends for screening-independent gynecologic cancers.^32,36,40,47,48^ Antunes et al. reported a decrease in newly diagnosed gynecological cancer patients in 2020 followed by an increased frequency of more aggressive histological types of endometrial and ovarian cancer in 2021.^35^ Knoll et al. reported a 45% decrease in newly diagnosed gynecological cancers during the 2020 lockdown periods followed by an increase in the proportion of symptomatic patients presenting upon re-opening compared to pre-COVID-19 periods in Austria.^111^ A retrospective single-center study reported a reduction in the number of gynecological patient visits after COVID-19 despite a relative increase in the overall number of patients and in the proportion of those with more symptomatic (including increase in abnormal bleeding complaints) and advanced disease.^131^ A retrospective, single-center study reported a larger percentage of endometrial cancer diagnoses following complaints of abnormal bleeding (92.06% vs. 67.02% pre-COVID-19).^132^ Garret and Seidman reported a reduction of surgically-staged endometrial cancer patients at presentation after COVID-19.^133^ These findings support the notion that changes in access to care over the COVID-19 period can impact both cancer rates as well as worsening of outcomes.

Delays in cancer diagnosis due to disruptions in screening or cancer care are of great concern as several retrospective analyses have shown that delays and other disruptions in oncological care occurring during the COVID-19 period were associated with worse outcomes in both gynecological and other cancers^51^ including stage migration, disease progression,^21,34^ and increased cancer mortality.^21,69,134,135^ Further research is required to assess the full impact of these delays on incidence, stage and survival of female cancers. These findings highlight the importance of carefully weighing the benefits of public measures imposed to slow the spread of a virus against the risk of negative cancer outcomes due to disruptions in cancer screening.

### Could changes in other factors contribute to changes in billing rates?

Studies show that lifestyle factors such as stress, fear and depression, lack of exercise and a more sedentary lifestyle can negatively impact cancer outcomes.^136–141^ Studies over the COVID-19 period suggest that fear, depression and anxiety particularly impacted females.^127,142–144^ Some have shown how strict lockdowns and fear-based responses adversely affected cancer care and have estimated substantial long-term impacts on female cancer outcomes.^38,116,145,146^ There have also been reports of women developing swollen lymph nodes^147–152^ and bleeding abnormalities (including changes in frequency, abnormal uterine bleeding and hormonal symptoms)^153–168^ following COVID-19 vaccination. In Ontario, guidance documents addressing lymphadenopathy following COVID-19 vaccination were developed for breast cancer screening and medical imaging sites and primary care providers.^169^ Menstrual irregularities have also been correlated with stress responses, COVID-19 infections and long-COVID.^112,155,161,162,167,170–173^ Given that lymphadenopathy and abnormal bleeding are part of the symptomology of female cancers^147–149,174,175^ and that OHIP billing likely reflects both suspected and confirmed cancers, it is possible that these symptoms may have confounded female cancer diagnoses contributing to the excess billing rates seen over the COVID-19 period.

To determine whether rates of bleeding irregularities changed in relation to OHIP billing prevalence rates for female cancers, we assessed pre-COVID-19 billing rates for disorders of menstruation (OHIP 626) and menopause, post-menopausal bleeding (OHIP 627) to rates during the COVID-19 period. We found that these billing codes which include bleeding irregularities followed a similar pattern as female cancers. Decreases of billing prevalence rates in 2020 relative to the pre-COVID-19 period were followed by a recovery in 2021-2022 with rates remaining above projected levels into 2022 (Figure 2). The comparable pattern of change in bleeding abnormality diagnostic codes further supports the notion that increases in bleeding symptoms may have contributed to a rise in suspected gynecologic cancer billing rates. Although the relationship between hormonal changes and endocrine-sensitive breast cancer is well-established,^176–179^ the degree to which hormonal changes likely underlying these bleeding abnormalities have impacted hormone-driven cancers remains unclear. Further analyses of cancer registry databases are warranted to determine the impact of public health measures and other medical or lifestyle factors on female cancer rates throughout and beyond the COVID-19 period.

**Figure 2.**
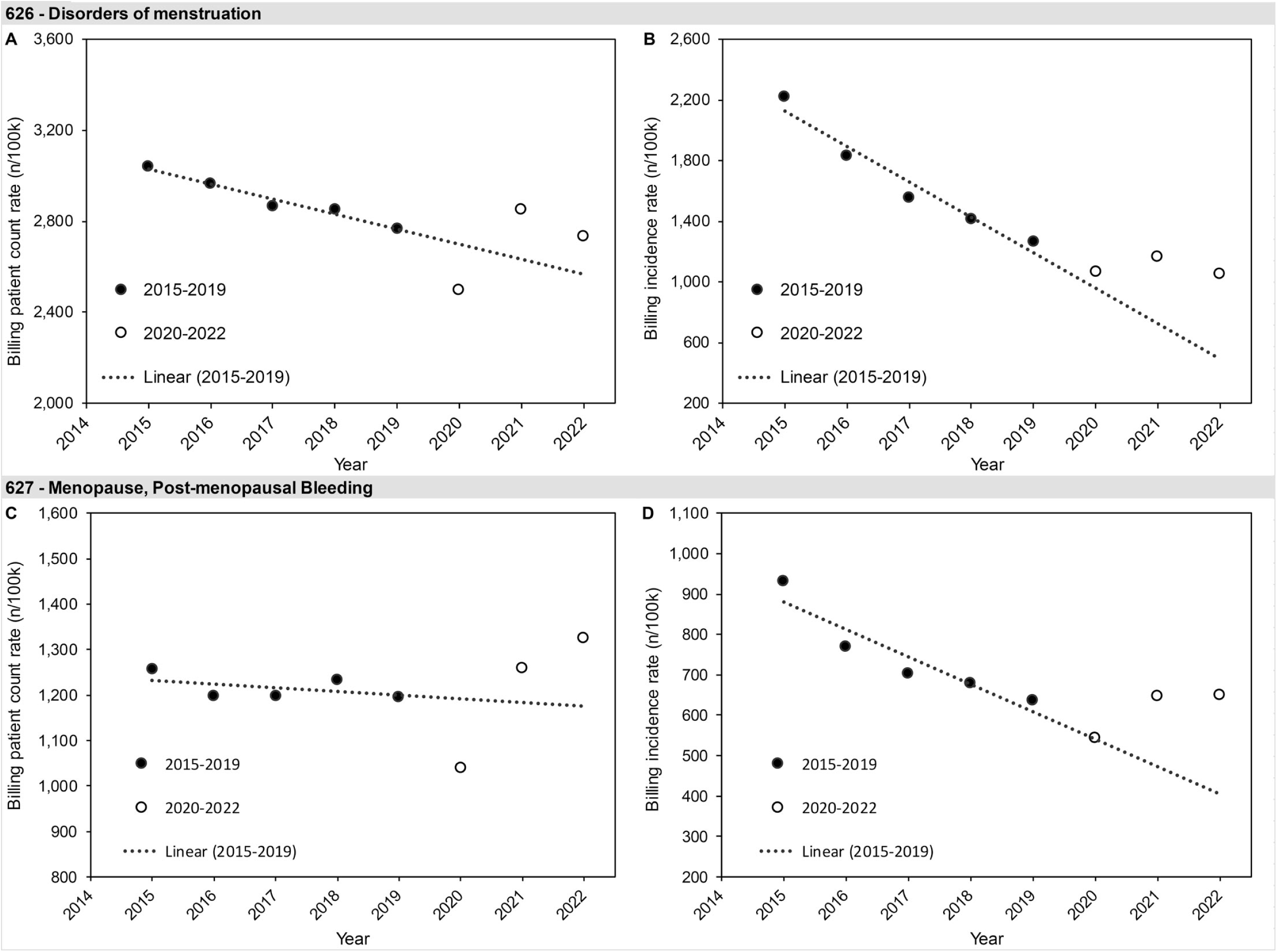
OHIP 2015-2022 billing patient count and incidence rates for disorders of menstruation (a,b) and menopause, post-menopausal bleeding diagnostic codes (c,d).

### What are the limitations of our analysis?

There are many limitations to our analysis, the primary one being its retrospective nature followed by a limited ability to formally assess the various changes to cancer care and how they may have affected cancer outcomes. Finally, the relationship between OHIP billing rates and cancer rates has yet to be elucidated and validated. Cancer registry analyses will be necessary to assess how patterns of care arising from public health measures may have impacted the incidence, stage and survival of women with female cancers.

#### Summary

Our analysis of OHIP billings rates for female cancers during the pandemic (2020-2022) compared to pre-pandemic levels (2015-2019) found an expected decrease in female cancer billing rates in 2020 followed by increases in 2021 and 2022, resulting in cumulative excess rates over the COVID-19 period, which were greatest for screening-dependent cancers. Disruptions to cancer screening and care in 2020 provide the most compelling explanation for the changes in billing patterns in 2020 and 2021 while increases in cancer-like symptoms in the COVID-19 period could contribute to the continued rise in billing rates through 2021 and into 2022. Our results highlight the importance of carefully weighing the effectiveness of public health measures designed to slow the spread of a virus against the risk of worsened cancer outcomes resulting from disruptions in screening and cancer care. Further research is required to assess the impact public health measures had on the incidence, stage and survival of women with female cancers.

## Declarations

### Ethics approval and consent to participate

Not applicable.

### Consent for publication

Not applicable.

### CRediT Author Contributions

**Alon D. Altman:** Conceptualization, Methodology, Writing-Reviewing and Editing **Christine Brezden-Masley:** Conceptualization, Writing-Reviewing and Editing **Nathalie LeVasseur:** Conceptualization, Writing-Reviewing and Editing **Ilidio Martins:** Methodology, Software, Formal Analysis, Writing-Original Draft, Writing-Reviewing and Editing, Visualization **Deanna McLeod:** Conceptualization, Methodology, Validation, Investigation, Resources, Writing-Reviewing and Editing, Supervision, Project Administration, Funding Acquisition **Amanda Selk:** Conceptualization, Methodology, Writing-Reviewing and Editing **Anna V. Tinker:** Conceptualization, Methodology, Writing-Reviewing and Editing

## Data Availability

All data produced in the present study are available upon reasonable request to the authors

## Acknowledgments

We would like to thank Karen Rucas and Dr. Susan Natsheh for their assistance in filling the FOI request and collecting the OHIP data and Akhil Padmanabhan from Kaleidoscope Strategic Inc. for research and editorial support.

## Funding

Funding for this article was provided through unrestricted educational grants from Eisai Limited (Canada) and The Society of Gynecologic Oncology of Canada (GOC). No discussion or viewing of review content was permitted with sponsors at any stage of manuscript development.

### Availability of data and materials

The datasets analyzed during the current study are available upon reasonable request.

