## Supplemental Figure 1 for "Changes in Female Cancer Diagnostic Billing Rates over the COVID-19 Period in the Ontario Health Insurance Plan"

Supplementary Materials

Figure S1. Overall billing prevalence and incidence rates for all diagnostic codes included in the dataset provided by OHIP

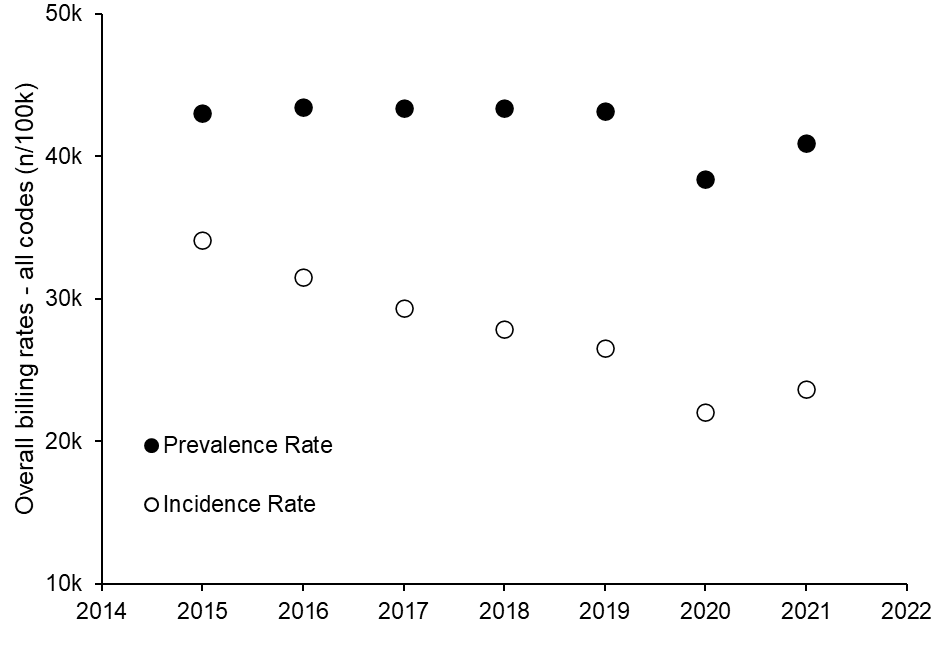

Abbreviations: n, number; OHIP, Ontario Health Insurance Plan
